# A combination of annual and nonannual forces drive respiratory disease in the tropics

**DOI:** 10.1101/2023.03.28.23287862

**Authors:** Fuhan Yang, Joseph L Servadio, Nguyen Thi Le Thanh, Ha Minh Lam, Marc Choisy, Pham Quang Thai, Tran Thi Nhu Thao, Nguyen Ha Thao Vy, Huynh Thi Phuong, Tran Dang Nguyen, Dong Thi Hoai Tam, Ephraim M Hanks, Ha Vinh, Ottar N Bjornstad, Nguyen Van Vinh Chau, Maciej F Boni

## Abstract

**Background:** It is well known that influenza and other respiratory viruses are wintertime-seasonal in temperate regions. However, respiratory disease seasonality in the tropics remains elusive. In this study, we aimed to characterize the seasonality of influenza-like illness (ILI) and influenza virus in Ho Chi Minh City (HCMC), Vietnam.

**Methods:** We monitored the daily number of ILI patients in 89 outpatient clinics from January 2010 to December 2019. We collected nasal swabs and tested for influenza from a subset of clinics from May 2012 to December 2019. We used spectral analysis to describe the periodicities in the system. We evaluated the contribution of these periodicities to predicting ILI and influenza patterns through lognormal and gamma hurdle models.

**Findings:** During ten years of community surveillance, 66,799 ILI reports were collected covering 2.9 million patient visits; 2604 nasal swabs were collected 559 of which were PCR-positive for influenza virus. Both annual and nonannual cycles were detected in the ILI time series, with the annual cycle showing 8.9% lower ILI activity (95% CI: 8.8%-9.0%) from February 24 to May 15. Nonannual cycles had substantial explanatory power for ILI trends (ΔAIC = 183) compared to all annual covariates (ΔAIC = 263). Near-annual signals were observed for PCR-confirmed influenza but were not consistent along in time or across influenza (sub)types.

**Interpretation:** Our study reveals a unique pattern of respiratory disease dynamics in a tropical setting influenced by both annual and nonannual drivers. Timing of vaccination campaigns and hospital capacity planning may require a complex forecasting approach.

**Funding:** National Institutes of Health, Wellcome Trust.

## Introduction

The epidemiology of influenza and other respiratory viruses is well understood in temperate regions and is characterized by predictable annual wintertime epidemics. However, the absence of winter in tropical regions makes the yearly patterns of influenza and influenza-like illness less seasonal and less predictable^1^. Thus far, it has been found that influenza in tropical regions shows lower variation in incidence as well as different periodicities temporally^2^ and spatially^3–6^, making it difficult to forecast periods of high incidence. Although some studies in tropical areas have shown associations between climate or environmental factors and influenza transmission^7–9^, this relationship remains elusive^1,10^ and caution is needed in interpreting these results and drawing inferences on a ‘tropical influenza season’.

There are two common shortcomings in many past analyses focused on influenza and ILI seasonality in tropical areas. First, it is not possible to generate robust evidence for seasonality using short time series with monthly data. Short time series may be unrepresentative of longer-term behaviors, and when combined with a monthly stratification of cases, provide low statistical power to determine when the respiratory disease seasons occurs if there is one. Second, it is not sufficient to base an analysis on associations between climate factors and ILI/influenza incidence when describing seasonality, as has been done previously^8,11,12^, because spurious associations will be common between an annually-structured set of climate factors and an event like an ILI epidemic that occurs historically about once per year. A determination of whether seasonality exists is needed first. Quantitative descriptions of long-term fine-scale time series are needed to accurately characterize the presence and pattern of seasonality in respiratory disease incidence.

Here we present the periodic signals detected from 10 years of daily influenza-like-illness (ILI) reports and 7.5 years of molecular surveillance for influenza virus, collected from a community mHealth syndromic surveillance study in Ho Chi Minh City, Vietnam. Using time series decomposition and regression models, we attempt to identify periodicities in the ILI and influenza time series, and to evaluate the explanatory power of these periodicities on both high-incidence and low-incidence periods of ILI and influenza.

## Methods

### Study Design

Starting in August 2009, the Oxford University Clinical Research Unit (OUCRU) in Ho Chi Minh City began recruiting community outpatient clinics to participate in a daily ILI reporting program by standard mobile phone SMS messaging. A total of 89 clinics were recruited during the first five years of the study and the data collection ended on December 31 2019. Clinicians or nurses in each clinic sent daily text messages to OUCRU reporting the total number of visits, the number of the patients that had ILI symptoms, and the number of hours that the clinic was open that day. The ECDC ILI definition was used: (1) sudden onset of symptoms within the past 3 or 4 days; (2) one or more of the following general symptoms (*i*) fever with axillary temperature above 37.5°C, (*ii*) malaise, (*iii*) headache, or (*iv*) myalgia; and (3) one or more of the following respiratory symptoms (*i*) cough, (*ii*) sore throat, or (*iii*) shortness of breath. The percentage of ILI patients among total outpatient visits per day (%ILI) in each clinic from January 1, 2010 to December 31, 2019 is used as the primary data type in the analysis. The %ILI time series for each clinic was detrended to an ILI *ζ*-score (“zeta” score) to remove long-term decreasing trends seen in seven clinics^13,14^ (see S1 Text). Daily data were detrended using the *ζ*-score for each clinic, and then the arithmetic mean among all clinics reporting that day was calculated to get an aggregated ILI time series. This aggregated all-clinic *ζ*-score was smoothed with a 7-day moving average to remove weekend effects.

Starting May 23 2012, 24 of the participating outpatient clinics agreed to participate in additional molecular influenza surveillance. Based on a randomized schedule, one clinic per week was assigned to collect naso-pharyngeal swab samples for influenza molecular confirmation by reverse transcription polymerase chain reaction (RT-PCR). Samples were subtyped to identify A/H1, A/H3, and influenza B. Counts of molecular samples and the number testing positive for each subtype were aggregated into 21-day windows to ensure sufficient samples in each window. A daily ILI+ time series was constructed as the product of the influenza positivity rate each day (this is constant for 21-day stretches) and the aggregate all-clinic daily ILI *ζ*-score. The daily ILI+ was then smoothed with a 7-day moving average.

### Statistical analysis

We used autocorrelation functions, discrete Fourier transforms, and wavelet analyses to identify periodic signals in the ILI *ζ*-score and ILI+ time series. We then used simple cyclic step functions (called “cycles” in equations below) to infer the magnitude and timing of periodic fluctuation in the time series (see S3 Text).

To identify the predictive ability of the inferred cycles in the ILI data, we regressed the ILI *ζ*-score using lognormal model on a range of potential predictors including the inferred cycles, 7-day lagged ILI *ζ*-score, twelve climate covariates, and a school-term indicator function (Eq.1).

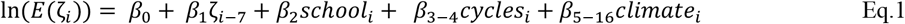

The 7-day autoregressive term was included because human-transmissible pathogen incidence time series are temporally autocorrelated. Climate data were collected from the NASA POWER Project^15^ and twelve climate covariates were included: temperature, absolute humidity, and rainfall, all lagged at 0, 1, 2, and 3 weeks, as all have been reported to be associated with ILI or influenza trends in previous studies^9^. School term was included because of the high transmissibility of ILI among children (see S4 Text).

In the molecular influenza time series, the statistical approach needs to account for an overrepresentation of zeroes in the ILI+ time series (about 9% of daily time points). We use a 2-step gamma hurdle model to regress ILI+ onto covariates.

The first step is a logistic model estimating the probability that influenza activity is present given the predictors (Eq.2). The second step is a gamma model estimating the magnitude of influenza activity conditioned on influenza activity being present on that day (Eq.3).

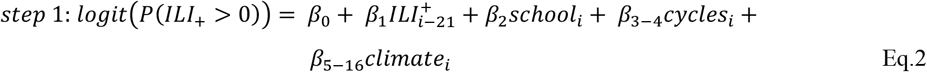

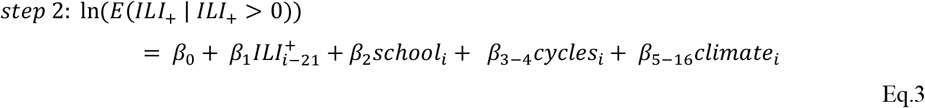

The autoregressive term of ILI+ is 21-day lagged ILI+. The inferred cycles were estimated from ILI+ data. All the other predictors remain the same as in ILI ***ζ***-score.

For both HCMC and US ILI data, we conducted stepwise AIC-based forward model selection to determine the predictors to include in the model. We compared the contributions of annual and nonannual cycles between HCMC and US ILI by decomposing the overall R-squared to each predictor (details in LMG in dominance analysis^16^ and S5 Text).

All analyses were conducted using R version 4.0.3. Wavelet analysis was done using WaveletComp package^17^. Gamma hurdle model was done using glmmTMB package^18^. R-squared decomposition was done using relaimpo package^19^.

## Results

From January 1, 2010 to December 31, 2019, 89 clinics were enrolled in the study. A total of 66,799 SMS text messages with ILI reports were sent covering 2,893,515 outpatient visits, 257,789 (8.9%) of which were patients meeting the clinical definition of ILI. Among the clinics, 33 were selected for analysis as they sent more than 300 reports with >50% of reports showing a non-zero number of ILI patients during the 10-year period. Among the included clinics, the median daily number of patients per clinic was 44 (IQR: 35 – 53), and the median of the daily number of patients per clinic meeting the definition of ILI was 4 (IQR: 3 - 6).

### Periodic signals in syndromic influenza data

The syndromic ILI *ζ*-score time series appears noisy (but is not white noise, p < 0.001, Box-Ljung test) and exhibits weak fluctuations with no visually discernible seasonality (Fig. 1), especially when compared to ILI patterns in temperate regions (Fig. S1). The absence of strong and regular seasonality is consistent with subtropical Hong Kong and tropical Singapore (Fig. S1). As the seasonal signals are not visually obvious, three separate analyses were used to determine the presence/absence of periodic signals in the data.

**Figure 1.**
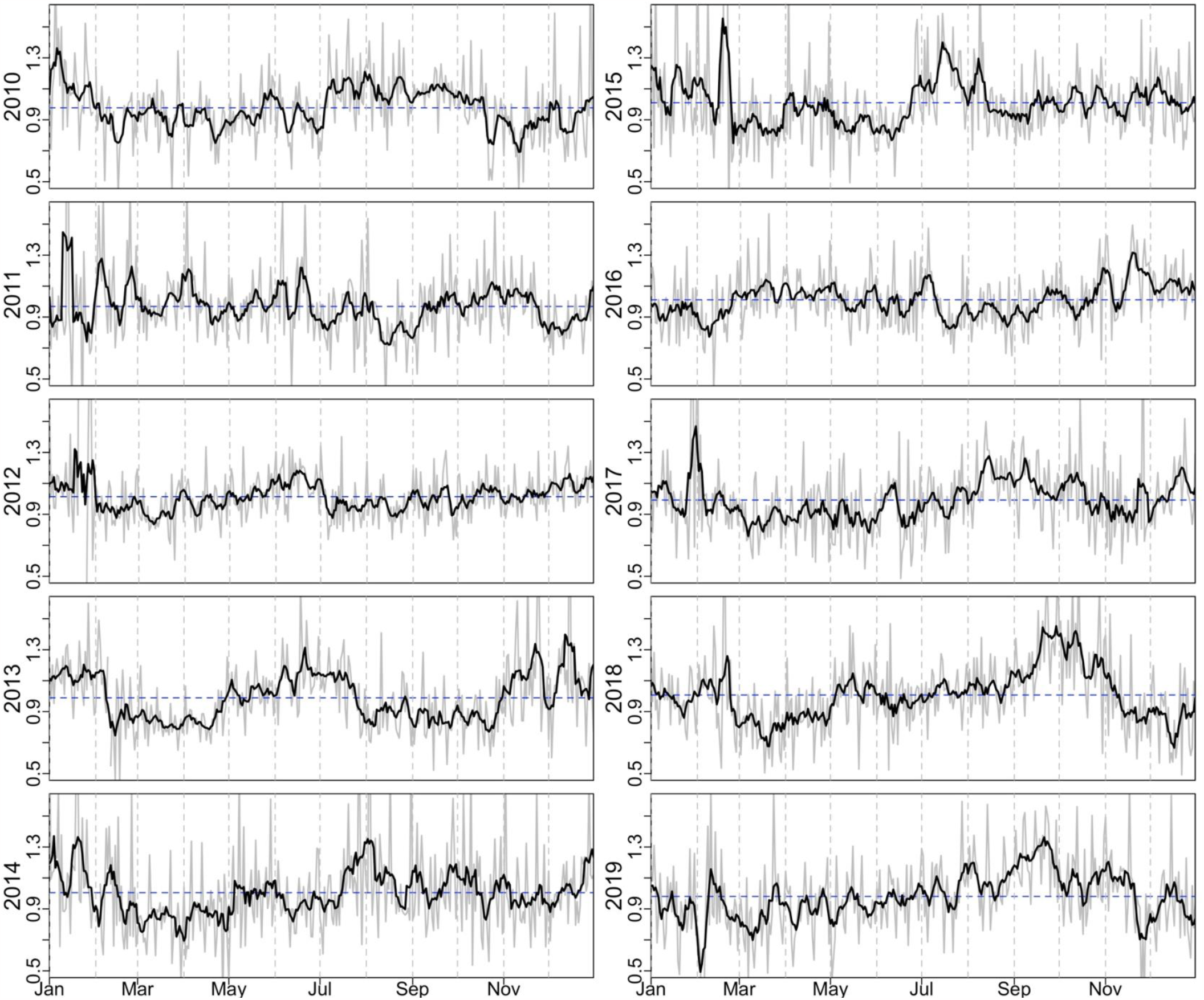
Daily ILI *ζ*-score (grey line) and 7-day smoothed ILI *ζ*-score (black line) from 2010-01-01 to 2019-12-31. The mean of ILI *ζ*-score in each year is shown as a blue horizontal dashed line.

Periodic signals detected by autocorrelation function (ACF) were weak in HCMC compared to temperate regions (Fig. S2), and they were not robust to the number of years included in the data. As in our previous analysis^13^, the first eight years of data collection showed a well-supported 203-day signal from 2010-2017 (Fig. 2A, top-left panels) and an annual signal appearing for most time periods. However, including all ten years of data from 2010-2019 showed a strong annual signal without a nonannual signal; this appears to be driven primarily by the high inter-year correlation in the ILI signals between the 2017/2018 and 2018/2019 (Pearson’s *ρ* are 0.325 and 0.568, respectively, Fig. S3). This shift from a primarily nonannual cycle to primarily annual cycle (Fig. 2B) is also observed in the wavelet analysis (Fig. S4). Statistical evidence (via the ACF) for both annual and nonannual cycles is robust to sub-setting the time series to shorter periods (Fig. 2B), except for the year 2019 which appears to have a singularly strong effect on the auto-correlation patterns. Discrete Fourier transform of the ILI *ζ*-score supports both a nonannual cycle (215 days) and an annual cycle (365 days) showing equally strong signals (Fig. S5).

**Figure. 2.**
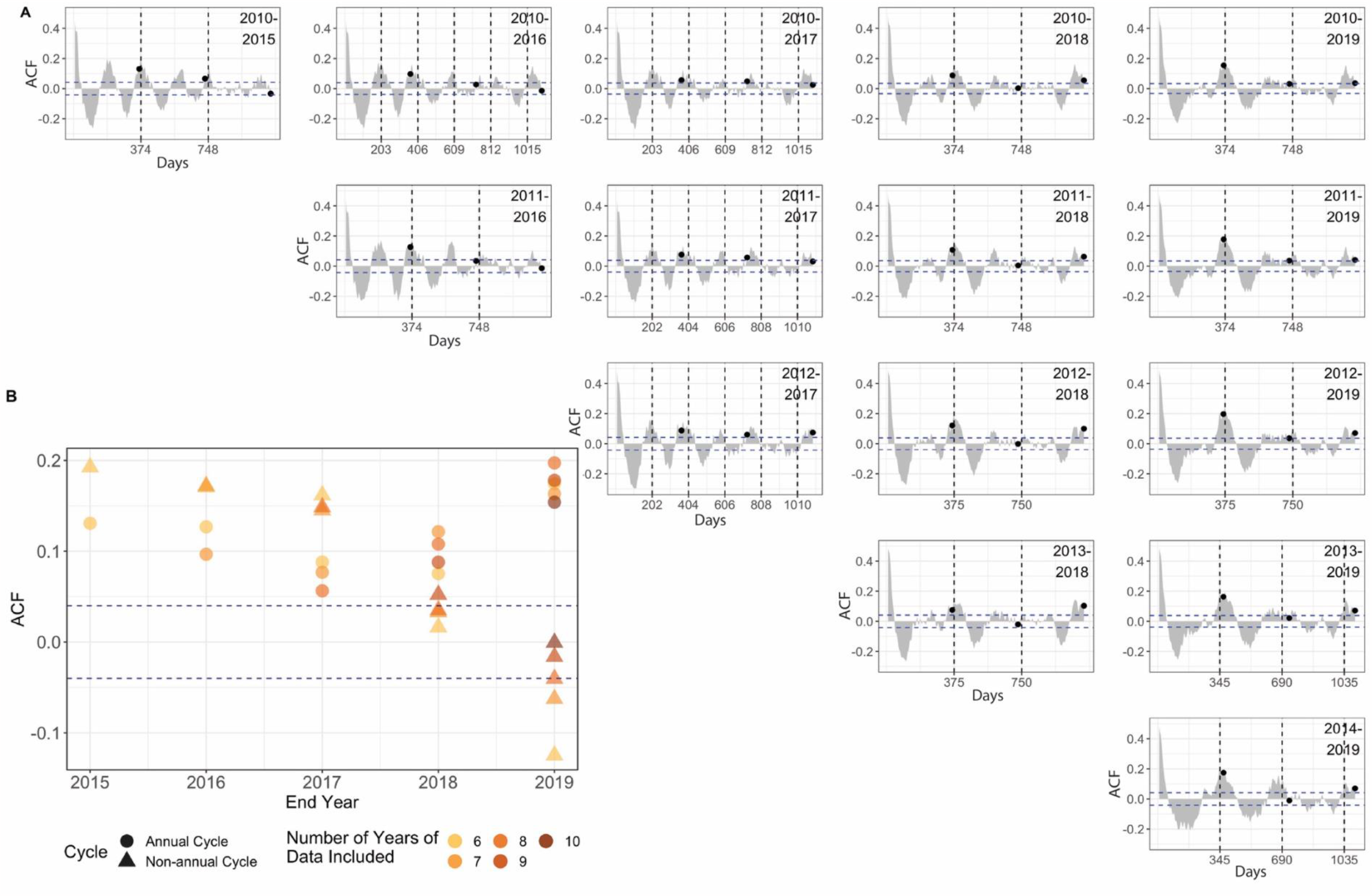
Nonannual and annual cycles in ILI *ζ*-score. (A) The Pearson autocorrelation function (ACF) of ILI *ζ*-score time series, split across different study periods. Horizontal blue dashed line labels the region where ACF is significantly different from 0 (p = 0.05). Vertical dashed line labels the day lag when the ACF is the highest between 150 to 450 days. Annual cycle is labeled with a black dot. Periods are inclusive so “2010-2015” spans six years. (B) The shift of ACF value from nonannual cycle to annual cycle. The *x*-axis denotes the last year included in the time series, and the *y*-axis shows the ACF value. ACF values of annual cycles (circles) and nonannual cycles (triangles) are shown. The nonannual cycle showed stronger signal at the through 2017.

Third, to describe the cycles quantitatively, a fit of a cyclic 2-step function to the ILI *ζ*-score selected 365 days and 210 days as the two periodicities most likely to explain the data (AIC = -5016 and AIC = -4931, respectively). For the annual cycle, the ILI *ζ*-score is 8.9% (95% CI: 8.8% - 9.0%) lower from February 24 (95% CI: Feb 24-25) to May 15 (95% CI: May 12-18), suggesting that respiratory disease seasonality in the tropics may manifest itself as a low season rather than a high season. For the 210-day cycle, the ILI *ζ*-score is 6.8% (95% CI: 6.6% - 7.0%) lower for a 104-day period of the cycle (see Fig. S6). In both cases, the difference in respiratory disease incidence between low and high season is small. The 8-step 365-day cycle (AIC=-5138) and the 5-step 210-day cycle (AIC = -5033) were selected from a varying number of steps and cycles (Fig. 3A) and were included in the regression analysis. After stepwise AIC-based model selection, annual and nonannual cycles are retained in the final regression along with the seven-day autoregressive term and various climate factors mainly related to humidity (Fig. 3C) (Table 1). The AIC difference when removing the nonannual cycle (ΔAIC = 183) was larger than when the annual cycle was removed (ΔAIC = 79), indicating that nonannual trends contain specific information for the ILI incidence pattern that is not contained in other predictors. The large contribution of the nonannual cycle to the model’s goodness-of-fit may signal the presence of certain nonannual epidemiological processes unique to tropical regions. The larger AIC difference (ΔAIC = 263) when removing all annual covariates suggests that ILI incidence showed a stronger annual pattern (data-derived and not necessarily climate-linked). The ΔAIC for all the climate factors alone is 50.

**Table 1.** Regression coefficients from the lognormal model of ILI *ζ*-score. The weather predictors were normalized using z-score standardization to have the same scale.

| **Coefficients** | **Estimate** | **95% Confidence interval** | **ΔAIC** |
| --- | --- | --- | --- |
| **Lognormal Model of ILI-ζ score** |  |  |  |
| **Intercept** | -1.193*** | [-1.348,-1.038] | NA |
| **7-day lagged ILI-ζ score** | 0.386*** | [0.357,0.416] | 611 |
| **Nonannual cycle** | 0.652*** | [0.559,0.745] | 183 |
| **Annual cycle** | 0.538*** | [0.422,0.655] | 79 |
| **Absolute humidity** | -0.02*** | [-0.028,-0.013] | 26 |
| **14-day lagged absolute humidity** | 0.019*** | [0.01,0.028] | 14 |
| **7-day lagged precipitation** | 0.013*** | [0.006,0.02] | 11 |
| **21-day lagged precipitation** | -0.009** | [-0.016,-0.002] | 4 |
| **21-day lagged temperature** | 0.006* | [0.001,0.01] | 3 |
| **21-day lagged absolute humidity** | 0.007 | [-0.002,0.016] | 0.6 |
| * p < 0.05<br>** p < 0.01<br>*** p < 0.001 |  |  |  |

**Figure 3.**
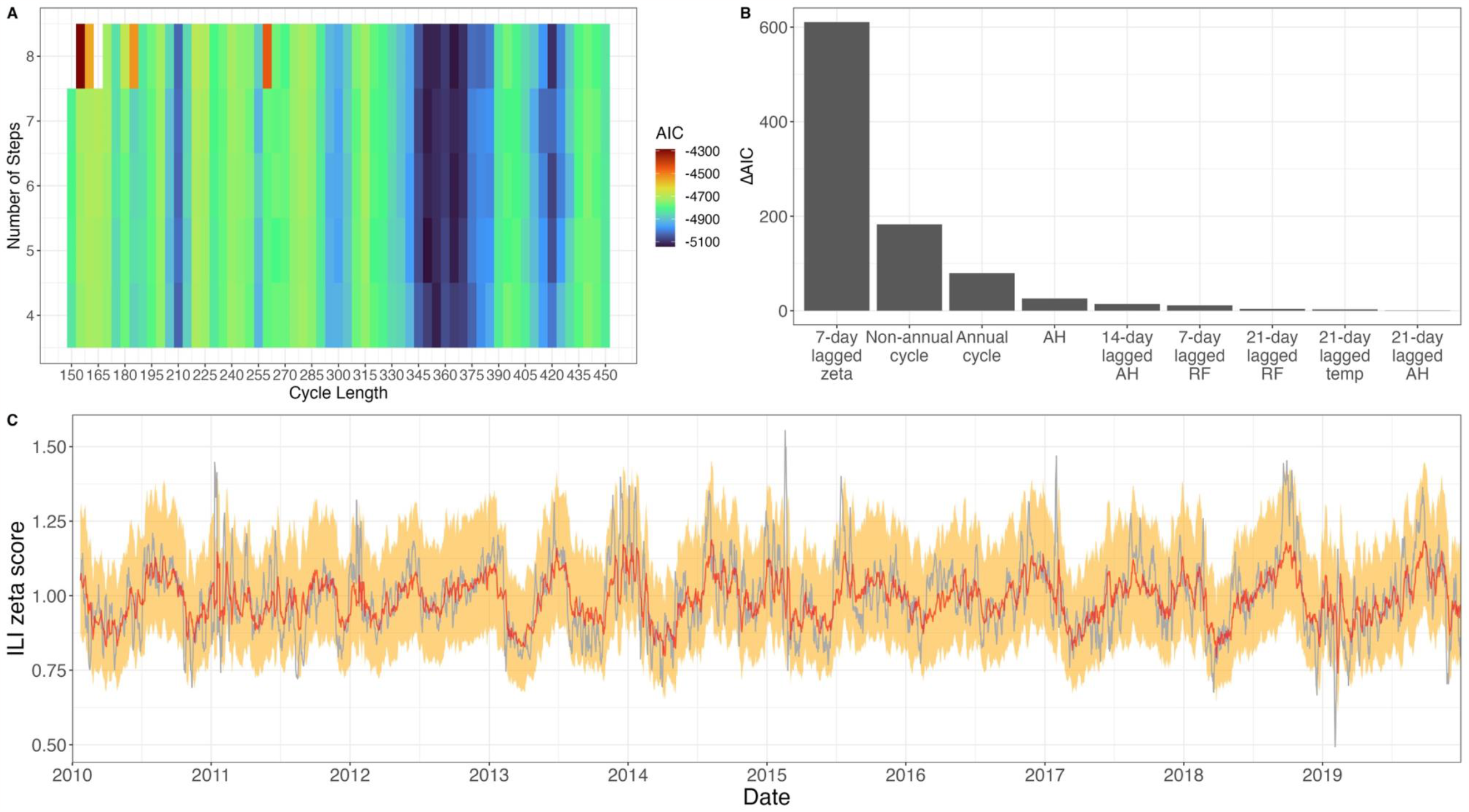
Nonannual and annual cycles in ILI *ζ*-score. (A) The AIC of cyclic step functions given ILI *ζ*-score among different number of steps and different cycle lengths. AIC is lowest when cycle is 210 days and close to annual. (B) Contribution of each predictor is calculated as the AIC difference when removing the predictor from the full model. (C) The predicted values of ILI *ζ*-score (red) and the 95% prediction intervals (orange) of the full model including both cycles and the observed ILI *ζ*-score (gray).

Critically, the nonannual ILI periodicities observed in HCMC are not present in temperate datasets that were processed with the same methods used for the HCMC data. Autocorrelation functions (Fig. S2) and wavelet analysis (Fig. S4) show strong peaks at one year with no signs of sub-annual periodicity – this is consistent across regional US data sets (10 HHS regions) and four European countries with time series longer than five years. Based on an R-squared decomposition^16^ from the regression models from HCMC and ten HHS regions, the nonannual inferred cycle explains around 15% variance of ILI *ζ*-score in the tropics compared to <1% of the variance in ten HHS regions (Fig. S7). Weaker annual signals were observed in HCMC than in temperate zones, but the nonannual signals were stronger.

### Periodic signals in influenza data

In the molecular surveillance component, a total of 2604 nasal swabs were collected from May 23, 2012 to December 31, 2019 of which 21.2% were positive for influenza. After subtyping, 6.3% were positive for influenza H1N1, 6.5% were positive for influenza H3N2, and 8.0% were positive for influenza B. There is no significant correlation between syndromic and virological data (Pearson’s *ρ* = -0.14, p-value = 0.11), suggesting co-circulation of many non-influenza respiratory pathogens.

To validate the molecular surveillance trends in our community-based study, we compared our ILI+ data to the ILI+ time series seen in hospital-based surveillance via the Vietnam’s National Influenza Sentinel Surveillance System^14,20,21^ in HCMC. The 186 weeks that overlapped during 2012 and 2015 between two time series showed similar circulation pattern of influenza and its subtypes, confirming the reporting consistency between our system and sentinel surveillance (Pearson’s correlation *ρ* = 0.568 (95% CI: 0.456–0.662) for overall influenza, *ρ* = 0.784 (95% CI: 0.719–0.836) for subtype A/H1N1, *ρ* = 0.706 (95% CI: 0.621–0.774) for subtype A/H3N2, and *ρ* = 0.523 (95% CI: 0.404–0.624) for influenza B; all *p* < 10^−4^, Fig. S8).

There is no conspicuous seasonality in influenza activity in HCMC (Fig. 4A). There appears to be a single influenza peak per year, with the autocorrelation function showing a maximum at 358 days (Fig. 4B) and the system showing ‘near-annual’ behavior rather than strictly annual behavior depending on how many years are included in the analysis (Fig. 4C). The discrete Fourier transform shows peaks at 324 and 417 days (Fig. 4D). A single influenza peak per year does not guarantee that peak timing is repeatable or consistent^14^. In addition, the peak timings and the periodic signals are not consistent across subtypes (Fig. 4A, Fig. S8, Fig. S9) suggesting a lack of climate or school-term influence on influenza circulation. The two-step function fit selected 330 days (AIC = -2414) and 385 days (AIC = -2405) as the dominant cycles in the overall ILI+ data, showing around a doubling of the ILI+ value in the high season (see S3 Text). The annual periodicity explains ILI+ less well (AIC = -2024) and shows weaker oscillation with a 55.1% (95% CI:[54.5 – 56.4]) increase from April 13 to July 18. The 8-step 330-day cycle (AIC = -2517) and 7-step 385-day cycle (AIC = -2491) were included in the final gamma-hurdle model (Table 2, Fig. 5C) after AIC forward model selection and are the most important predictors explaining the occurrence and the magnitude of influenza activity (Fig. 5B). The multiple near-annual periodicities in ILI+ suggest a period with high influenza activity that keeps shifting every year, a hypothesis that requires further testing.

**Table 2.** The coefficients from the gamma-hurdle model of ILI+. The weather predictors were normalized using z-score standardization to have the same scale.

| Coefficients | Estimate | 95% Confidence Interval | ΔAIC |
| --- | --- | --- | --- |
| **Logistic Model of ILI+** |  |  |  |
| Intercept | -3.069*** | [-4.055, -2.118] | NA |
| 21-day lagged ILI+ | 8.060*** | [6.383, 9.862] | 118 |
| 385-day cycle | 12.213*** | [9.547, 15.004] | 92 |
| Temperature | -1.007*** | [-1.227, -0.794] | 92 |
| School term | 1.921*** | [1.361, 2.489] | 43 |
| 21-day lagged rainfall | -0.715*** | [-0.997, -0.426] | 20 |
| 7-day lagged absolute humidity | -0.537*** | [-0.817, -0.268] | 14 |
| Rainfall | 0.591** | [0.216, 0.991] | 8 |
| 330-day cycle | 3.532** | [1.140, 6.040] | 7 |
| 7-day lagged rainfall | 0.481* | [0.121, 0.872] | 5 |
| **Gamma Model of ILI+(link = ‘log’)** |  |  |  |
| Intercept | -3.148*** | [-3.266, -3.029] | NA |
| 330-day cycle | 2.501*** | [2.205, 2.799] | 254 |
| 385-day cycle | 3.086*** | [2.721, 3.451] | 251 |
| 21-day lagged ILI+ | 0.975*** | [0.825, 1.125] | 154 |
| 21-day lagged temperature | 0.127*** | [0.080, 0.173] | 25 |
| School term | 0.189*** | [0.120,0.259] | 25 |
| Absolute humidity | -0.075*** | [-0.112, -0.039] | 14 |
| Rainfall | 0.089*** | [0.043, 0.137] | 11 |
| 21-day lagged rainfall | 0.070** | [0.022, 0.119] | 6 |
| Temperature | -0.056* | [-0.102, -0.009] | 3 |
| 7-day lagged rainfall | 0.053* | [0.008, 0.099] | 3 |
| * **p < 0.05**<br>** **p < 0.01**<br>*** **p < 0.001** |  |  |  |

**Figure 4.**
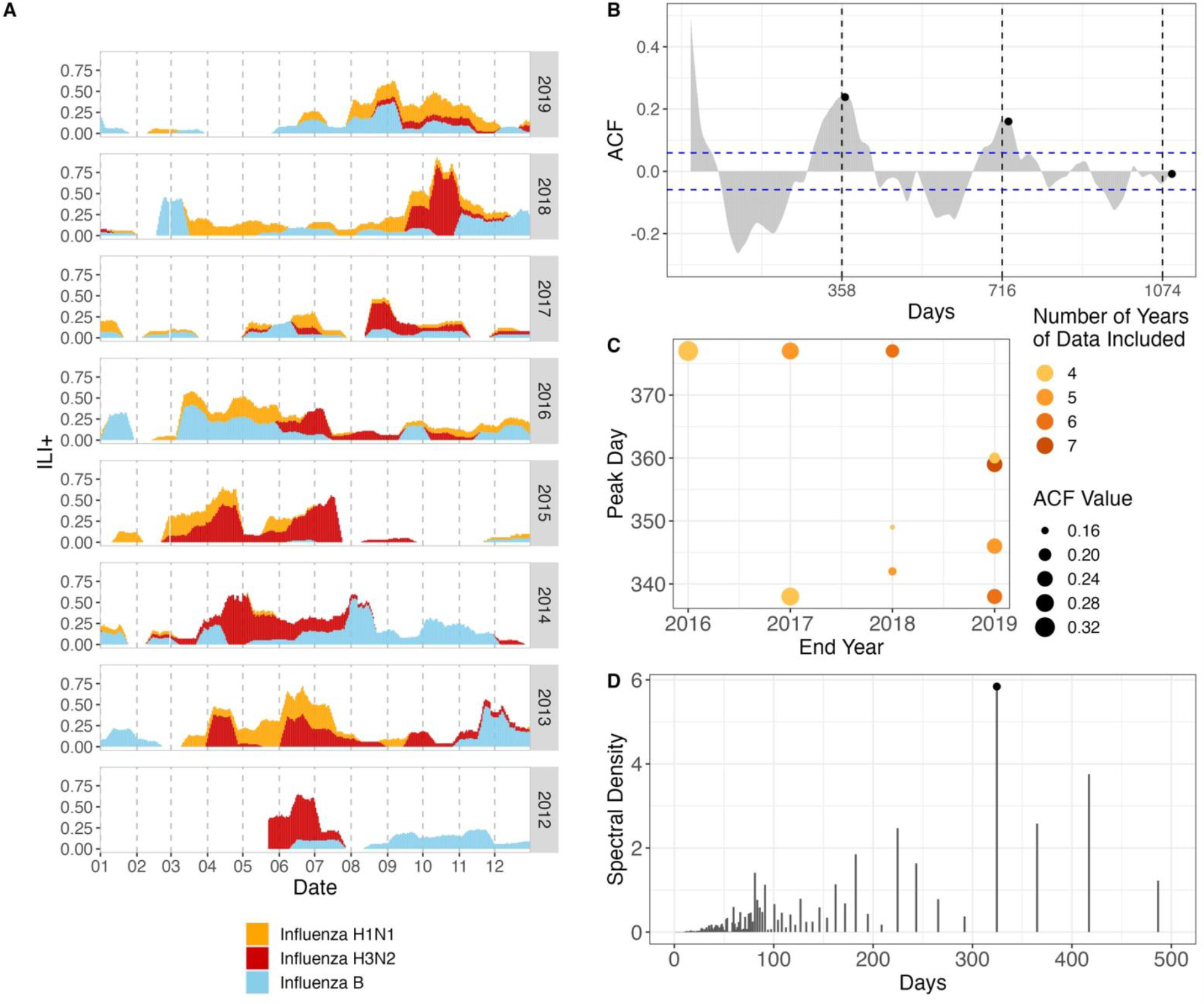
ILI+ time series and periodic signals. (A) The 7-day smoothed overall ILI+, stacked by subtypes. (B) ACF plot of the entire time series of overall ILI+. Vertical dashed line labels show ACF peaks at 358-day lag and the subsequent cycles. Black points show annual cycles. (C) The peak lag of ACF varied between 338 and 377 days in varying length of included ILI+ time series. (D) Discrete Fourier transform of the entire time series of overall ILI+. Black circle labels the dominant 324-day cycle.

**Figure 5.**
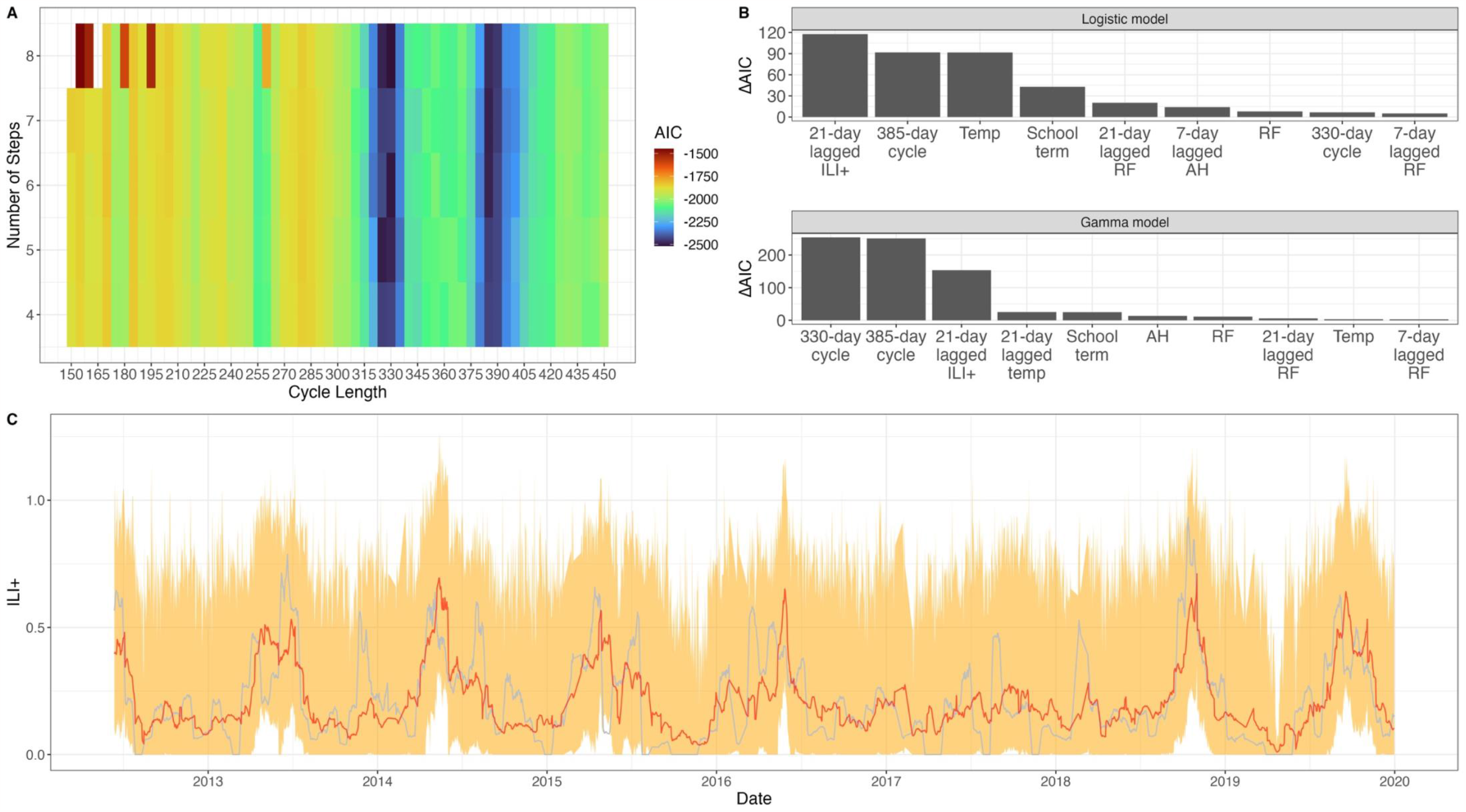
Cycles in ILI+.(A) AIC heatmap for cycle length and number of steps allowed in each cycle. Near-annual cycle lengths (330-day and 385-day) have the best fits. (B) AIC contribution of each predictor in gamma hurdle model, measured as the AIC difference when removing the predictor from the final model. (C) The predicted ILI+ (red) with 95% prediction interval (orange) is shown with observed ILI+ (gray).

## Discussion

Our community-led syndromic mHealth study was designed to remove barriers to enrollment, simplify reporting, and encourage long-term consistent participation in order to generate a syndromic data stream comparable to the ‘big data’ epidemiological outputs that began to be assembled at the beginning of last decade. Our purpose here was to generate a medium-sized data stream of ∼10^5^ data points where each data point was traceable back to a physician diagnosis. The proof that a ‘medium data’ approach can work at this scale is the validation of our community study’s influenza time series against Vietnam’s national sentinel surveillance system, showing the same incidence patterns for influenza A/H3N2, A/H1N1, and influenza B over a 186-week period. Additionally, the daily reporting in this study provides a unique level of resolution in identifying annual or cyclic patterns of disease incidence.

### Mechanisms

There is unlikely to be a specific climate effect driving ILI or influenza dynamics. In our data set, climate covariates tend to have low explanatory power (Fig. 3B, Fig. 5B). Different subtypes peak at different times of year, suggesting an improbable link between climate factors and the particular influenza subtype they are influencing.

Seasonality explanations in temperate regions are clearer: the environmental factors associated with viral survival and transmissibility and human movement/contact behaviors all experience abrupt changes during winter. With no winter-forcing in the tropics, we may observe the disease dynamics and their associated cycles driven by other factors. That is, the observed periodic signals in our study may be indicative of the natural internal clockwork of the dynamics of respiratory viruses^22^. Another possibility is that, we observed transient dynamics fluctuating around endemic equilibrium perturbed by stochastic events (super-spreading events, traveling waves), which will create cyclic behaviors^23^.

A lack of correlation between ILI incidence and PCR-confirmed influenza incidence indicates that a broader interpretation of all respiratory virus circulation is necessary to understand the mechanisms of a more complex system with many viruses competing for resources, excluding other viruses through short-term immune interactions, and driving a pattern of incidence in humans that is visible to us as a series of low peaks and long shallow troughs of ILI. A long-term cohort with a weekly panel of molecular diagnostics would be the right starting point for describing the interactions among a large group of respiratory viruses.

Finally, a reassessment of the definitions of ILI or flu season is needed in characterizing respiratory virus circulation in the tropics. The “outbreak” or “epidemic” designation is commonly used in temperate regions to describe a more-than-5-fold increase in ILI activity, a criterion that could not be used in the tropics. Instead, year-round persistence with 9% lower activity during a 13-week period suggests that ILI transmission could experience a short low season in the tropics, inferred as late-February to mid-May in HCMC. Retooling mechanistic models to identify periods of low transmission may be the next step in understanding the long-term effects of this particular epidemiological driver. Identification of the driving forces of respiratory virus dynamics in the tropics is still very much an open question.

### Limitations

Difference in health-care access between high-income and low-income settings may lead to discrepancies in the populations being represented in syndromic surveillance reporting. For this reason, temperate and tropical ILI trends may not be directly comparable as general-population measures. Our study showed a high correlation between community surveillance and hospital-based surveillance (Fig. S7), but comparison between Vietnam and Hong Kong or Singapore would be necessary to determine if health-care access has any effect on regional patterns of respiratory disease transmission in Southeast Asia^24,25^.

Due to the nature of the data smoothing for the purpose of the analysis presented (S1 Text), lag periods in the regression models were constrained to be multiples of seven days for ILI and 21 days for ILI+. These were based on using seven-day smoothing to remove weekend effects and 21-day smoothing to have sufficient sample sizes for diagnostic test results. This limits the ability to include the factors that have an effect on ILI or influenza at a time lag shorter than seven days. For example, the results that the climate factors in the regression models carried low influence on the overall model fit may be associated with the exclusion of short-term climatic effects, which may represent important mechanisms in influenza transmission.

The model fits shown from both the models for ILI and ILI+ in Ho Chi Minh City showed close fits to the data (Fig. 3C, Fig. 5C). However, the goodness of fit may be impacted by using the previously-fit step functions as covariates as these covariates were derived from the data. This potentially leads to overfitting in the models, where a function of the observed ILI(+) values was used as a predictor of ILI(+). However, given the definitions of the step functions, the predicted values from the models are likely similar to those that used the timings of the steps from the step functions as a set of binary predictors since fitted values from the step function were constant throughout each step.

### Long-term outlook

The methods and results in this study can be extended into a forecasting framework in order to predict future peaks or incidence of influenza in Ho Chi Minh City. Similar work using statistical models to produce short-term forecasts of infectious disease burden has been applied to respiratory viruses as well as other non-respiratory human communicable diseases and vector-borne diseases^26–29^. In the context of this study, it is shown that the regression models predict ILI and ILI+ with high accuracy (Fig. 3C, Fig. 5C), suggesting the potential of high predictive power for future burden. Extending the methods used in this study to produce a forecasting model is a natural extension of the current study. While regular forecasts of incidence may prove difficult due to the model’s setup to predict detrended data rather than actual incidence values, the forecasted trajectories would prove useful in identifying periods of relatively high ILI or ILI+ incidence.

If the approaches presented here lead to successful forecasts of ILI and ILI+ peaks, this may help inform prevention measures for influenza such as vaccination, public health messaging, and preparation for hospital capacity. Influenza vaccination coverage in Vietnam is currently low^30^, though efforts to increase vaccination among healthcare workers have been introduced in recent years. Noting the lack of strong annual seasons is important for designing vaccine campaigns in Ho Chi Minh City because there is little evidence to show that there is an optimal time for administering vaccines. Likewise, public health messaging and preparation in medical system for influenza and other respiratory diseases have the potential for substantial improvement if influenza and ILI patterns in Vietnam and other tropical regions can be better understood and forecast more accurately.

## Data Availability

All data produced are available online at https://github.com/bonilab/tropicalflu-03FL-10years-communityILI-HCMC.

## Data Availability

All the data and code are available at https://github.com/bonilab/tropicalflu-03FL-10years-communityILI-HCMC.

## Author Contributions

Conceptualization – FY, JLS, MFB

Methodology – FY, JLS, HML, EMH

Software – TDN

Validation – FY, MC, PQT

Formal Analysis – FY

Investigation – TTNT, NHTV, HTP

Resources – N/A

Data Curation – FY, PQT, MC

Writing (Original Draft) – FY

Writing (Review and Editing) – FY, JLS, MFB

Visualization – FY

Supervision – MFB, ONB, JLS, MC, EMH

Project Administration – MFB, NTLT, DTHT, HV NVVC

Funding Acquisition – MFB

