## Supplementary materials for "A combination of annual and nonannual forces drive respiratory disease in the tropics"

*S1 Text: Time series detrending*

There are 7 clinics showing a long-term downtrend in %ILI. To remove the long-term trend and get a stationary time series, we detrended the %ILI by dividing each daily value by a 365-day moving average centered at that day for each clinic. We refer to this transformation as a ζ-score as it is related to an exponentiated z-score. This removes trends longer than 365 days and preserves the ratios between sequential daily reporting numbers. For influenza, we calculated daily ILI+ as the product of the mean of the daily ILI ζ-score across all the clinics and the influenza positivity rate on that day to get a time series representing influenza incidence. Then we applied a 7-day moving average to smooth both the ILI ζ-score and ILI+ to filter out short-term noise. The 7-day moving smoothed ILI ζ-score and ILI+ are used for subsequent analysis.

*S2 Text: ILI data from other surveillance systems*

To compare ILI trends in HCMC with other regions, ILI data from the United States, four European countries (Belgium, France, Greece, Netherlands), Singapore, and Hong Kong were collected. The ILI data from the United States was downloaded from ILINet (US CDC). The weekly weighted %ILI from October 5 1997 to December 29 2019 from 10 HHS (Health and Human Services) regions were downloaded and then used as the overall ILI time series in the United States. The ILI data from Portugal, Belgium, Greece, and Netherlands were all downloaded from ECDC. The weekly number of patients showing ILI symptoms per 100,000 population from October 5 2015 to December 30 2019 was used. The ILI data from France were downloaded from France Sentinelles network https://www.sentiweb.fr/?lang=en. The weekly number of patients showing ILI symptoms per 100,000 population from November 3 1984 to December 28 2019 was used. The ILI data from Singapore were parsed from the Weekly Bulletin of the Singapore Ministry of Health. The average daily number of patients seeking treatment for ILI per week was available from January 4 2015 to December 28 2019. ILI data from Hong Kong were downloaded from the Department of Health in Hong Kong, from December 29 2013 to December 28 2019. All ILI data were converted into an ILI ζ-score time series with the same methods above.

*S3 Text: Cyclic step function*

We used simple step functions with $k$ steps in a periodic cycle of length $c$. The function values and breakpoints were estimated by minimizing the Akaike Information Criterion (AIC) using normally distributed errors. We maximized likelihood using the Nelder-Mead algorithm through the step function with cycle lengths $c$ ranged from 150 to 450 days, and the number of steps $k$ ranged from 2 to 8. AIC would be penalized as positive infinite if the length of a step is shorter than 15 days. The estimated parameters were selected from 100 optimized step functions with randomly selected initial parameters to ensure a global optimum was found. 95% confidence intervals were obtained via likelihood profiling.

For the ILI $\zeta$ -score, a final 8-step function was chosen for the annual cycle, and a final 5-step function was chosen for the nonannual cycle. For the ILI+, a final 8-step function was chosen for 330-day cycle, and a final 7-step function was chosen for 385-day cycle.

For ILI+, annual seasonality is inconsistent across subtypes. Subtype H3N2 shows the strongest annual periodicity, while H1N1 and influenza B appear to show no regular cyclic pattern (Fig. S8). The two-step function fit selected 330 days (AIC = -2414) and 385 days (AIC = -2405) as the dominant cycles in the overall ILI+ data, exhibiting a 105.4% (95% CI:[104.8 – 106.2]) increase during an 85-day high period (330-day inferred cycle) and a 106.2% (95% CI:[105.0 – 107.2]) increase during a 230-day high period (385-day inferred cycle, details in Fig. S6).

*S4 Text: Regression Covariates*

We collected climate data from the NASA POWER project[15]. Based on the criterion to only include climate factors that were reported to be biologically or epidemiologically associated with ILI transmission, we collected daily temperature, absolute humidity, and precipitation. We also included the 1-week, 2-week, 3-week lagged version of all climate variables because the effect of climate on ILI and influenza can be delayed. Absolute humidity was calculated using temperature and relative humidity:

$$AH = \frac{0.611 \times e^{\frac{17.502 \times T}{240.97+T}} \times 2.168 \times RH}{273.15+T} \quad \text{Eq.4}$$

Each climate predictor was normalized using the mean and the standard deviation of the predictor time series and then smoothed using 7-day moving average. The school-term categorical variable is one from August 15 to June 1, when schools in HCMC are in session and zero otherwise.

*S5 Text: Predictor importance*

In multiple linear regression, the importance of each predictor in regression was measured as the averaged R-squared difference when adding the predictor in all the possible subsets of predictors in the model. It is also referred as LMG in dominance analysis[19], defined as:

$$LMG(x_k) = \frac{1}{p}\sum_{i=0}^{p-1}\left(\sum_{\substack{S=\{x_1, x_2, \dots x_p | x_k\} \\ n(S)=i}} \frac{R^2(x_k \cup S) - R^2(S)}{\binom{p-1}{i}}\right) \qquad \text{Eq.5}$$

where p is the number of predictors, S is all the subsets of predictors except $x_k$. We compared the predictor importance between the US ILI **ζ**-score and the ILI **ζ**-score in Ho Chi Minh City. For US ILI **ζ**-score, we started with fitting the step function given the cycle length is between 21 weeks and 64 weeks. The annual cycle was the fitted step function given the cycle length is 52 weeks, and the nonannual cycle was the best fitted step function out of the step functions when their cycle length is between 27 and 31 weeks (corresponding to the interval 190-220 days of the nonannual cycle in our ILI **ζ**-score). The 52-week cycle was always the best fitted step function in the ILI-**ζ** score in all the HHS regions. Next, we respectively regressed the ILI **ζ**-score in all the HHS regions using the same predictors as in the data in Ho Chi Minh City, including lagged climate factors from each HHS region, school term, 7-day lagged ILI **ζ**-score, and fitted cycles. We calculated the predictor importance from all the selected models using AIC-based selection, and we compared with the predictor importance of the model of ILI **ζ**-score in Ho Chi Minh City.

**Supplementary Figures**

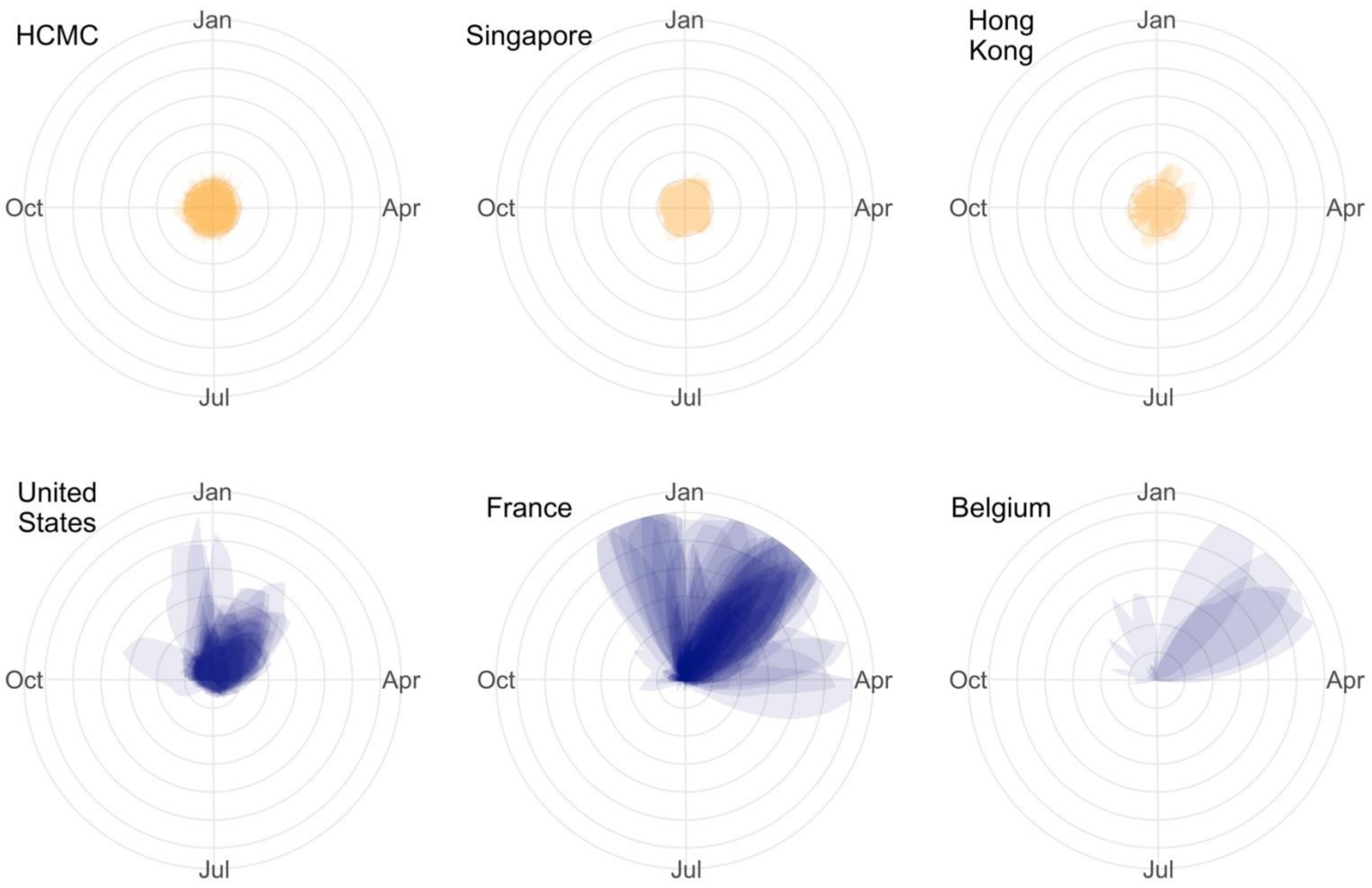


Figure S1. The circular plots of weekly ILI ζ -score from HCMC, Singapore, Hong Kong, the United States, France, and Belgium (data sources: S2 Text). The orange color indicates tropical or subtropical regions, the blue color indicates temperate regions. The data from each year was visualized in a circular time scale. The range of each plot is [0,6]. The weekly ILI ζ -score from HCMC is calculated by averaging the daily ILI ζ -score within one week.

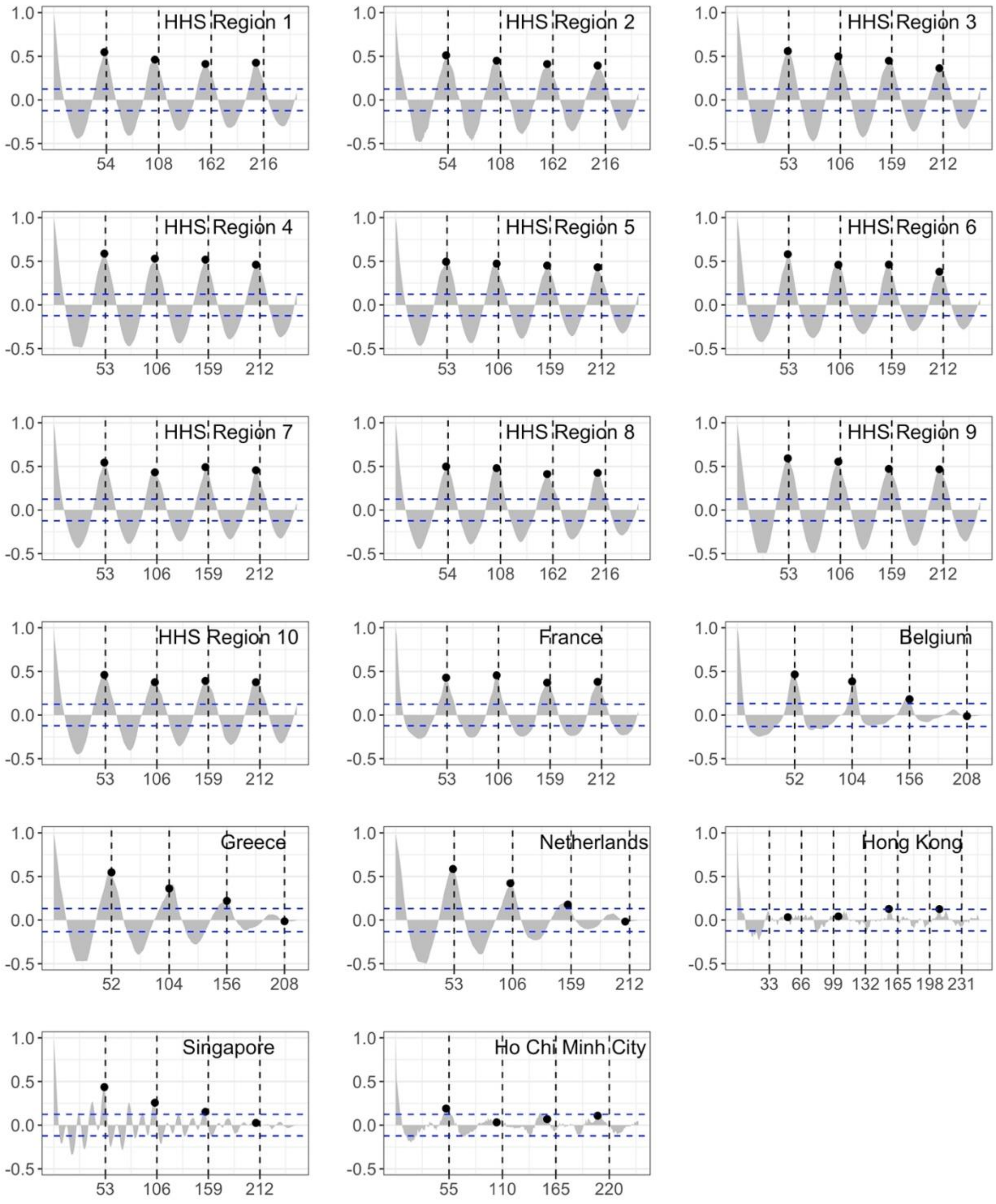


Figure S2. ACF plots of the weekly ILI ζ -score from locations in temperate regions and tropical regions (data sources: S2 Text). Compared to temperate regions, the ILI and in HCMC exhibit weak annual seasonality. The weak seasonality is also observed in Hong Kong and Singapore.

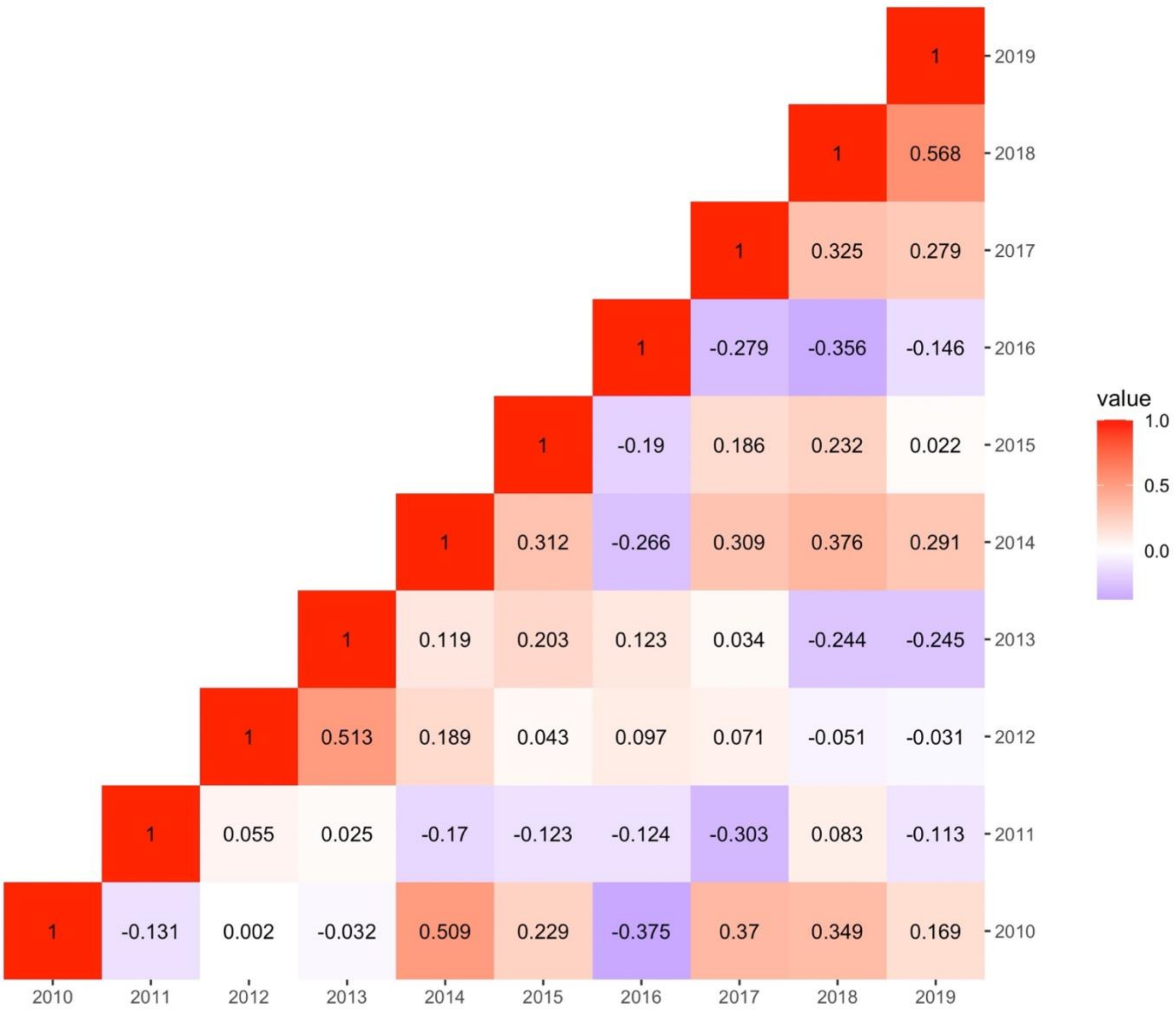


Figure S3 Pearson's correlation of ILI ζ -score between different years from 2010 to 2019. Notice the high correlation between 2018 and 2019.

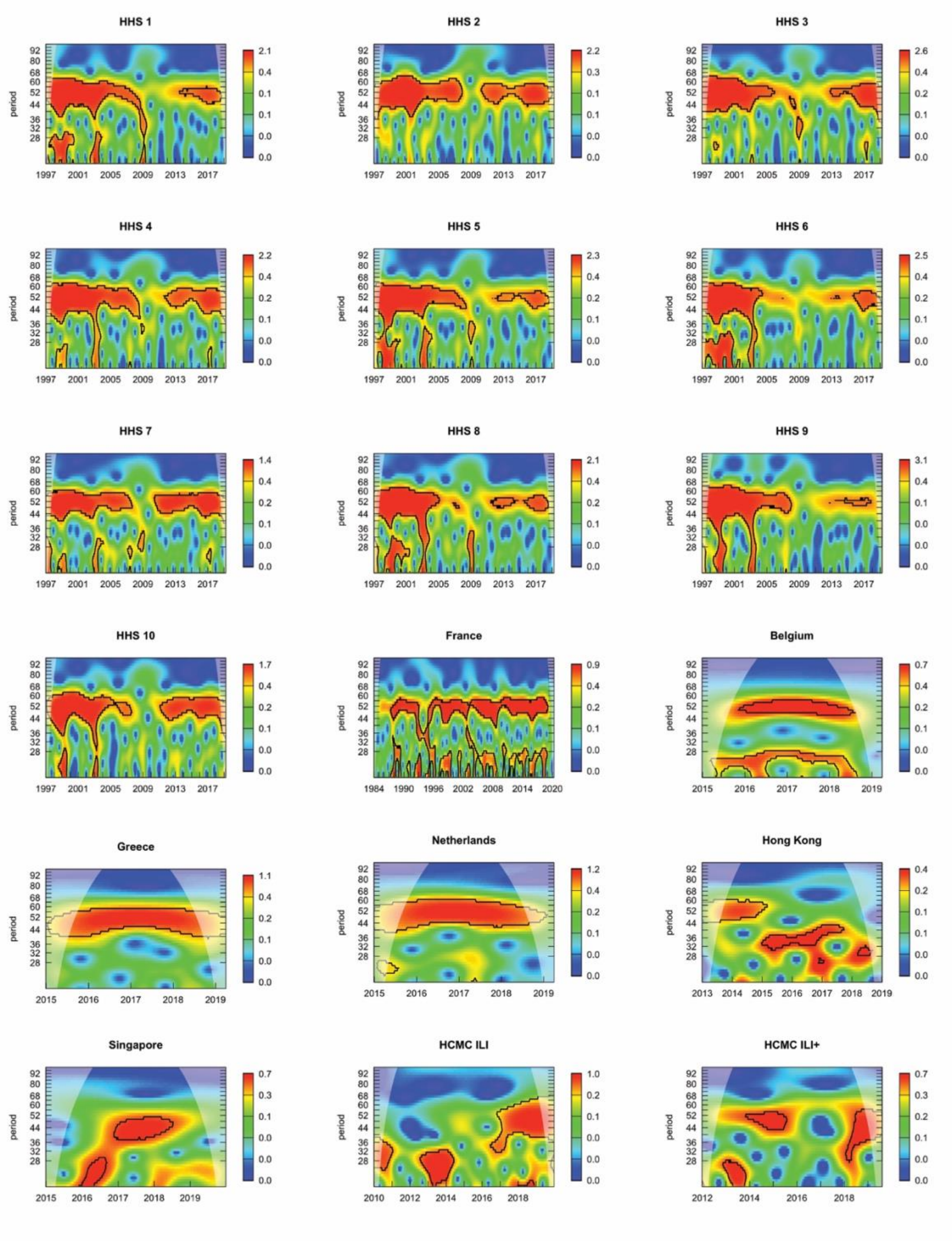


Figure S4 Wavelet transform of the ILI ζ -score in ten HHS regions from the United States, four European countries including France, Belgium, Greece, and Netherlands, and subtropical city Hong Kong, and tropical country Singapore, along with the and ILI+ from HCMC. Continuous annual seasonality (52 weeks) is only observed in temperate regions. The disruption of annual seasonality in the HHS regions was caused by 2009 H1N1 pandemic.

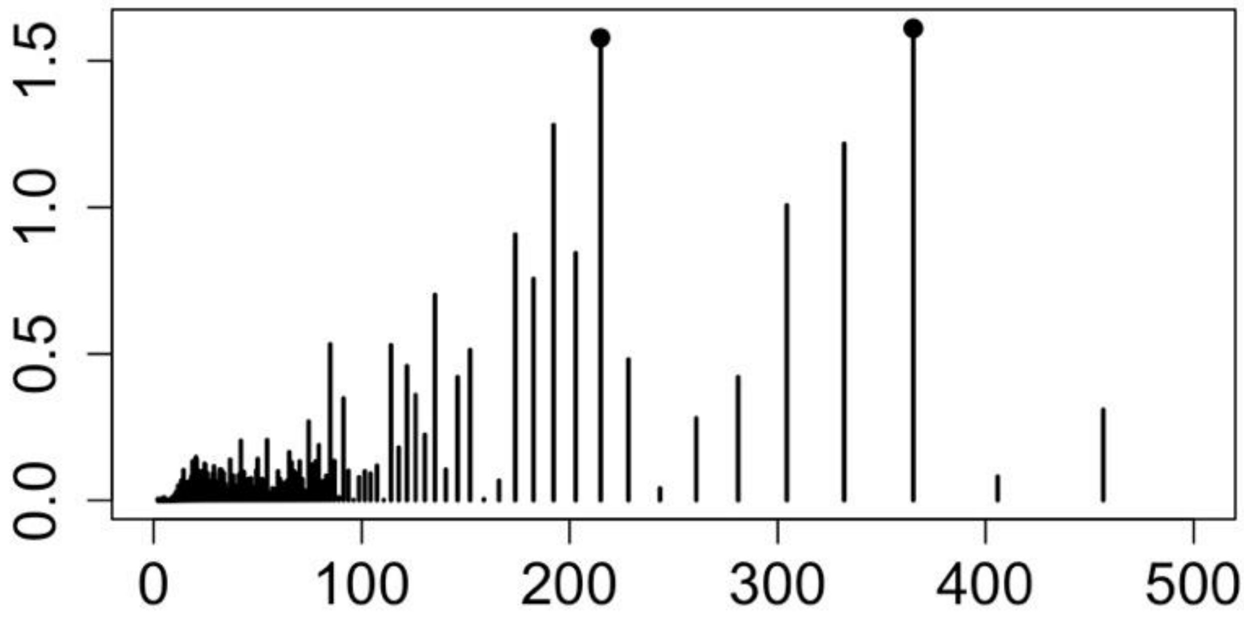


Figure S5 Discrete Fourier transform of ILI ζ -score. The signal of 215-day cycle and 365-day cycle are equivalently strong in the ILI ζ -score.

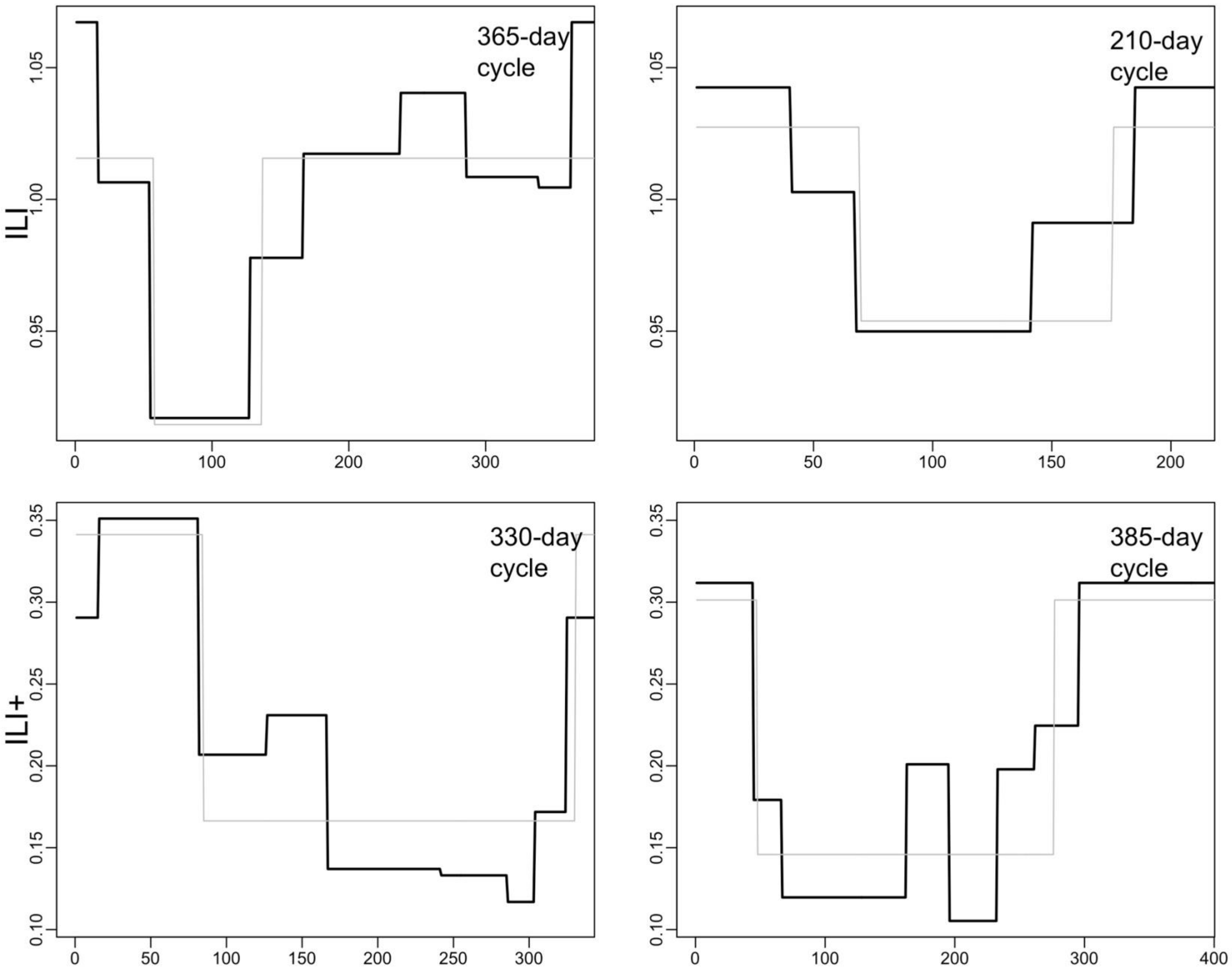


Figure S6. The inferred cycles of ILI ζ -score and ILI+. The grey lines indicate the 2-step cycles. The black lines indicate the multiple-step cycles.

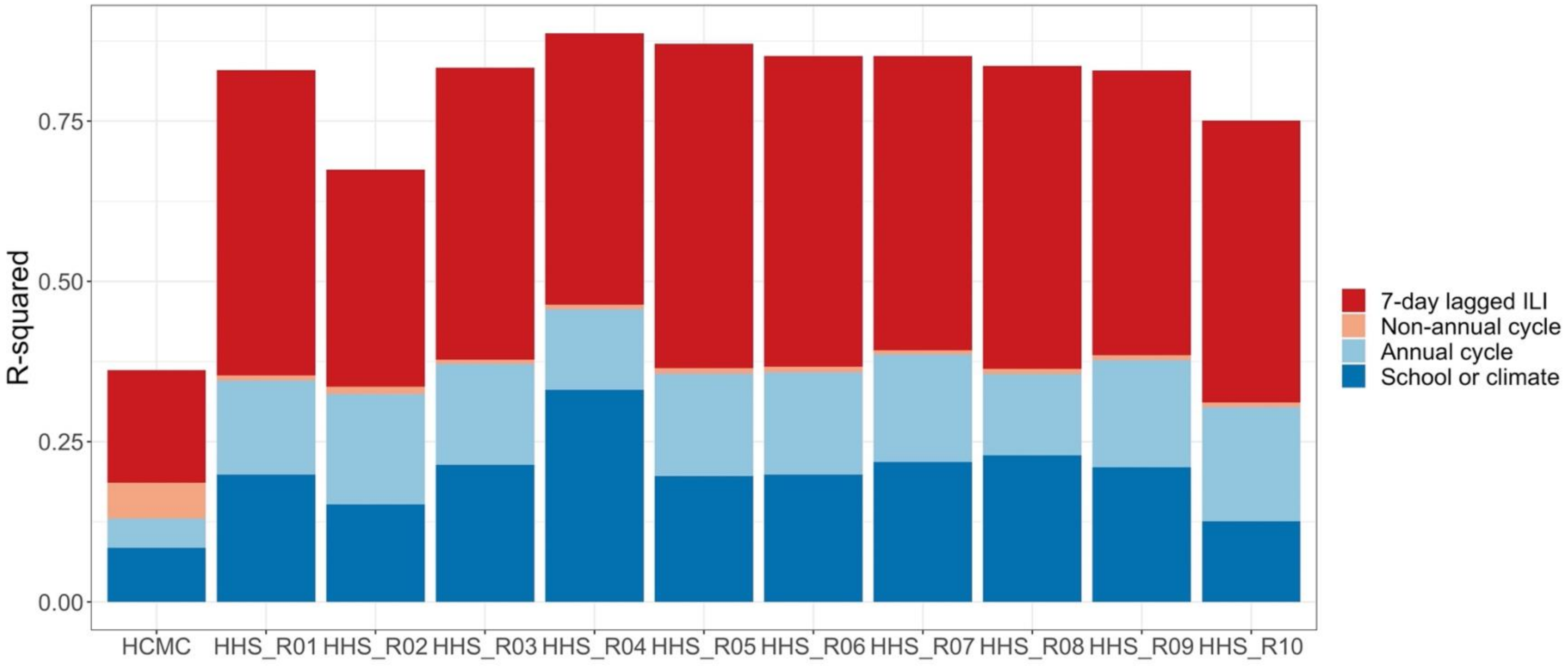


Figure S7. R-squared partitioned to the predictors with school term and climatic factors combined from the models of the ILI ζ -score from HCMC and ten HHS regions from the United States.

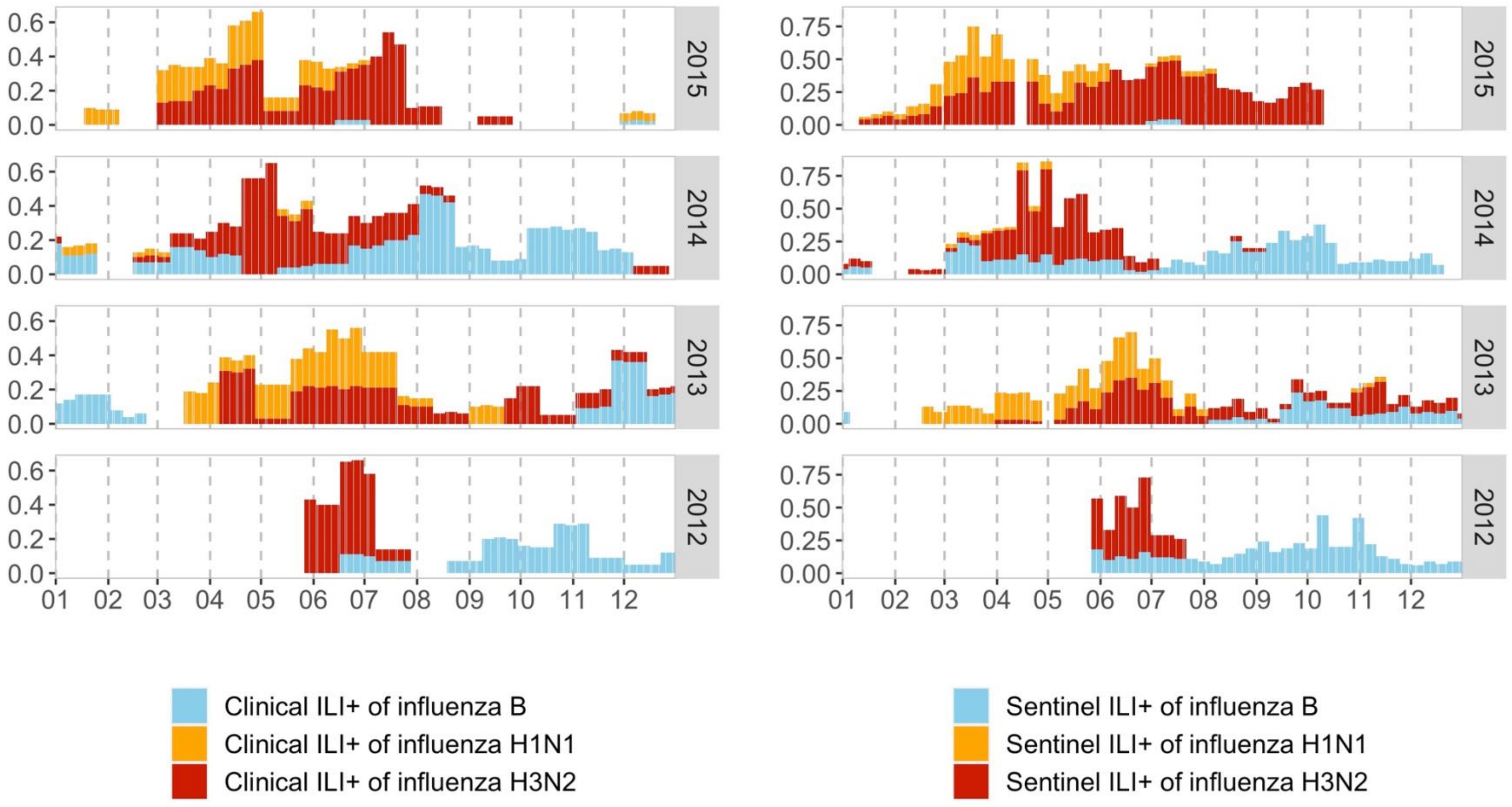


Figure S8. ILI+ from our surveillance system (Left) and Hospital for Tropical Diseases in Ho Chi Min City (Right). Weekly ILI+ is calculated to be comparable to the hospital's weekly ILI+. During the period when two time series are both available, the ILI+ shows similar trend.

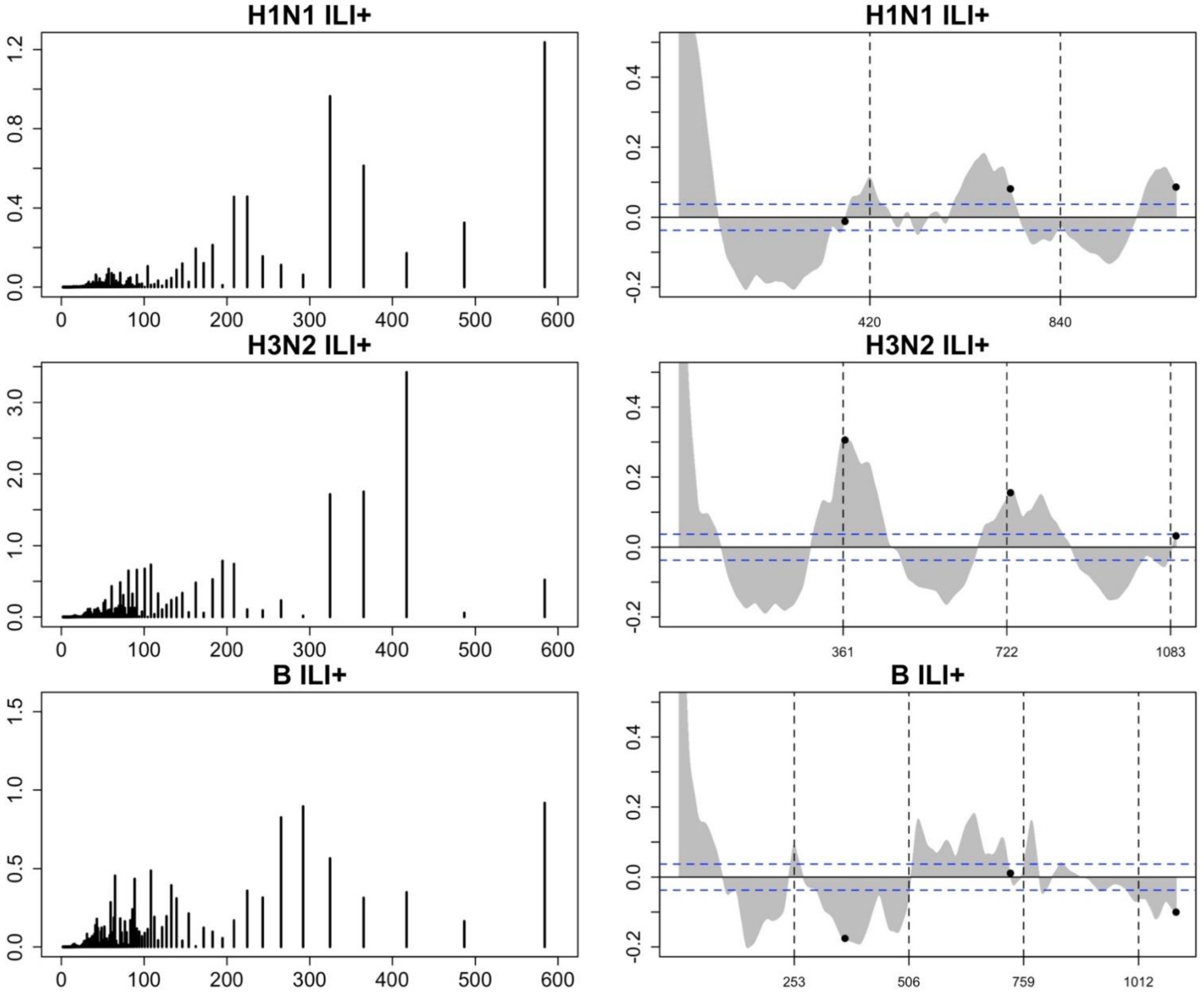


Figure S9. Discrete Fourier transform and ACF plots of subtype ILI+. Based on the spectral density and the ACF coefficients, H3N2 ILI+ showed the strongest annual periodic signal, while H1N1 ILI+ and influenza B ILI+ showed irregular pattern in the absence of annual seasonality.
